# The economic burden of cervical cancer on women in Uganda: a cross-sectional study

**DOI:** 10.1101/2023.10.26.23296906

**Authors:** Hallie Dau, Esther Nankya, Priscilla Naguti, Miriam Basemera, Beth A. Payne, Marianne Vidler, Joel Singer, Avery McNair, Maryam AboMoslim, Laurie Smith, Jackson Orem, Carolyn Nakisige, Gina Ogilvie

## Abstract

**PURPOSE:** There is limited research on how a cervical cancer diagnosis financially impacts women and their families in Uganda, including the burden of out-of-pocket costs. This is important to understand because, in addition to being economic providers, women are primary caregivers. The objective of this analysis is to describe the economic impact of cervical cancer treatment, including how this differs by socio-economic status (SES) in Uganda.

**METHODS:** We conducted a cross-sectional study from September 19, 2022 to January 17, 2023. Women were recruited from the Uganda Cancer Institute and Jinja Regional Referral Hospital, and were eligible for this study if they were ≥ of 18 years and being treated for cervical cancer. Participants completed a 45-minute survey which included questions about their out-of-pocket costs, unpaid labor, and changes in their family’s economic situation. A wealth index was constructed from the participants ownership of household items to determine their SES. Descriptive statistics were reported.

**RESULTS:** Of the 338 participants who completed the survey, 183 were from the lower SES. Women from the lower SES were significantly more likely to be older, have ≤ primary school education, and have a more advanced stage of cervical cancer. Both groups of women reported paying out-of-pocket for cervical cancer care (higher SES 95.5%, lower SES 92.3%). Only 15/338 participants stopped treatment because they could not afford it. Women of a lower SES were significantly more likely to report borrowing money (p-value=0.004) and selling possessions to pay for cancer care (p-value=0.006). With regards to unpaid labor, women from both groups reported a decrease in the amount of time that they spent caring for their children since their cervical cancer diagnosis.

**CONCLUSION:** Regardless of their SES, women in Uganda incur out-of-pocket costs related to their cervical cancer treatment. However, there are inequities as women from the lower SES groups were more likely to borrow funds to afford treatment. Alternative payment models and further financial support could help alleviate the financial burden of cervical cancer of care on women and families in Uganda.

## INTRODUCTION

The burden of cancer is rapidly growing in Africa. It is estimated that by 2040, the number of new cancer cases in Africa will increase by 37% and cancer deaths will increase by almost 40%. Moreover, the burden of cervical cancer in Africa is expected to rise from 117,000 new cases in 2020 to 164,000 in 2030.[1] This is significant because cervical cancer is an entirely preventable disease as a result of vaccination and screening tools. Currently, sub-Saharan African health systems are largely ill-equipped to handle the steadily increasing number of cancer cases. Much of the cancer care response to date has been reactive rather than proactive, with more investment in treatment than prevention.[2] Challenges in cancer care within the continent can be attributed to insufficient training of healthcare staff, absence of diagnostic facilities, weak coordination systems, and, most critically, insufficient funding.[2] This in turn has made it difficult for women to access high-quality cancer screening and treatment services.

Few countries in sub-Saharan Africa currently offer universal healthcare.[3] In Uganda, user fees from national healthcare costs were eliminated in 2001, which resulted in an increase in seeking care at government-sponsored healthcare facilities.[4] However, it has widely been reported that individuals still often pay out-of-pocket for care at these facilities as not all costs are in fact universally covered.[5-8] While well-known anecdotally, there is little research on how a cancer diagnosis and subsequent treatment can financially impact individuals and their families in Uganda. Of the literature available, a large portion, including that specific to cervical cancer, is qualitative in design.[9-11] In 2022, Germans et al. published a qualitative study on the socio-economic burden of cervical cancer on women in Uganda which largely focused on the financial burden. They found that a cervical cancer diagnosis leads to economic disruptions and can further exasperate a family’s financial security. However, this sample was limited to rural participants seeking care at hospice centers and did not report on indirect financial outcomes such as reducing necessary household expenditures to pay for treatment costs and changes in unpaid labor (e.g., child minding).[10]

It is important to understand the economic impact of a woman’s cancer treatment as it can have both far reaching consequences. Economic impacts are defined as any change to an individual’s financial status and can be direct (e.g., costs of medical treatment) or indirect (e.g., loss of work). A 2021 study by Ngcamphalala et al. in Eswatini found that loss of productivity (i.e., loss of wages) due to premature death from cervical cancer is a significant burden of indirect costs.[12] Moreover, previous research has shown that a quarter of patients at the Uganda Cancer Institute (UCI) have lost their jobs as a result of their cancer diagnosis.[13] However, this research is limited in its applicability here as it does not specifically speak about cervical cancer. In Uganda, like many parts of the world, women spend one and a half more time on childcare when compared to men.[14] This means that when women are diagnosed with cervical cancer, unpaid labor such as child minding, needs to also be considered along with traditional measures of impact such as lost wages and unemployment when speaking about the overall economic impact.

Currently, there are only two studies in low- and middle-income countries that quantify the direct and indirect costs of cervical cancer treatments.[15, 16] However, as they are limited to two countries, they provide a limited understanding of the global burden. The first study by Hailu et al. in Ethiopia measured both direct costs such as consultation fees and indirect costs such as lost wages. The study team concluded that cervical cancer has a significant financial burden on women diagnosed as women often have to pay for direct costs such as treatments, transportation, and medication. Moreover, they reported that women lost an average of 119 working days during the course of their treatment.[15] The second, Singh et al. from India, focused primarily on direct costs such as treatment fees and reported similar findings.[16] Not only is it a tragedy that women and their families are experiencing economic hardship as a result of their cervical cancer treatment, but moreover this is stemming from a preventable disease. It is important to quantify the economic burden of cervical cancer because it can be used to provide evidence to invest in the expansion of preventative programs, such as cervical cancer screening, which can reduce the long-term incidence of cervical cancer and ultimately reduce the economic burden on women. As such, additional research is needed in more countries to highlight the economic burden of cervical cancer. The objective of this analysis is to describe the economic impact of cervical cancer treatment, including how this differs by socio-economic status in Uganda.

## METHODS

### Data collection

This analysis is part of a larger exploratory cross-sectional study which aims to understand the social and economic impacts of cervical cancer on women and their children in Uganda. We recruited women from September 19, 2022 to January 17, 2023 using convenience sampling at the gynecology-oncologic department at UCI in Kampala and the cancer unit at Jinja Regional Referral Hospital (JRRH). Oncology nurses trained in data collection procedures recruited women and orally administered a 45-minute cross-sectional survey on electronic tablets using REDCap software [17]. As not everyone who participated in the study was literate, we chose to orally administer to survey for equity and to reduce bias. Women were eligible for the study if they were (i) aged ≥18 years with a histopathologic diagnosis of cervical cancer; (ii) being treated at the UCI in Kampala or JRRH cancer clinic for cervical cancer; and (iii) able to provide consent to participate in the study in English, Luganda, Lusoga, Luo, or Runyankole.

### Survey tool

Out-of-pocket medical costs were informed by two existing surveys. The first was a cross-sectional study from Nepal by Khatiwoda et al. which measured the cost of cancer care.[18] The second was a cross-sectional study from Uganda on pediatric surgical expenditure by Yap et al.[5] Women in the study were asked to note what forms of medical care they had paid out-of-pocket for (e.g., chemotherapy, medication, bed, consult). Following, women were asked to provide the total amount that they had paid out-of-pocket since their cancer diagnosis. In addition to their cancer care, women were also asked how much they paid out-of-pocket for the indirect costs of transportation, housing, and food in the past month. These variables were recategorized to be a yes or no question which answered whether or not women paid out-of-pocket for any of these expenses. Women who did not provide a monetary amount were recategorized as missing.

Women were asked if anyone in their family received any type of support for their treatment or care. Check all options included cash support, waiver support (e.g., housing voucher), in-kind support, none, or do not know. Women were also asked if they needed any food, transportation, or accommodation support. Additionally, women were asked if they stopped or refused any cancer care because of the costs. If they answered yes, they were prompted to note which treatment(s).

The survey asked women if they had to reduce household expenditures to help pay for their cervical cancer treatment. Select all response options included borrowed money from family, asked for charity/begged, sold possessions (e.g., land, car, furniture), moved to more affordable housing, withdrew child from school, reduced food consumed, changed transportation type, cut down on other needs, no, and do not know.

The survey included questions on the woman’s employment. One question asked women if they took any time off work to attend the clinic today. If applicable, women were also asked if any of the adults in their family (excluding themselves) either left their employment, reduced the number of hours worked, increased the number of hours worked, or missed work because of their cervical cancer treatment. Answers included no, yes, not applicable, do not know, and refused. Only women who provided a yes or no answer were reported in this analysis. All other answers were recategorized to be missing.

In addition to the monetary impact that their cervical cancer treatment had on their family, women were also asked about unpaid labor. Women were asked how the amount of time had changed (increased, decreased, stayed the same not applicable, do not know) that they spent on certain household chores since their cervical cancer diagnosis. Questions included buying food, caring for children, cooking food, cleaning the house, retrieving water, assisting children with work, and washing clothes.

Demographic variables included in this analysis were as follows: participants age, marital status, education, economic status, number of children in their household, occupation, economic status, time since diagnosis, cancer stage, and cancer treatment received. Additional variables collected related to SES were informed by Rutstein[19] and were as follows: home floor type, cooking fuel type, toilet type, residence type, and electricity in the home were considered for the wealth index as well as ownership of a smartphone, basic phone, computer, sewing machine, bicycle, car, motorcycle, fridge, and sofa. Smartphone and basic phone were recategorized as “phone” and car and moto were recategorized as “vehicle”.

### Analysis

Counts and frequencies were used to construct all descriptive statistics. Bivariate data was analyzed using chi-square and fisher-exacts tests. P-values ≤ 0.05 were considered significant. Records with more than 75% of missing data were removed from the analysis. All analyses were conducted using R 4.2.3.[20]

Principal component analysis was used to construct the wealth index.[21, 22] All considered variables were first recoded to be binary. The variables floor material type,[23] cooking fuel type,[24] toilet type,[25] drinking water source,[25, 26] and residence type.[19] were categorized as either improved (yes) or unimproved (no) based on global standards. The remaining variables were categorized as either own (yes) or do not own (no). Following, all of the asset variables were reviewed and those in which under 5% or over 95% of the sample indicated yes, were removed.[27] A complete case analysis was used as there was a small percentage (3.7%) of rows with missing data.[22] The R package “psych” was used to conduct the principle component analysis.[28-30] The varimax rotation was used with one component for extraction was included in the final index.[22, 29, 30] In all, nine participants were excluded from the wealth index due to insufficient data for computation. The final index included the variables: improved floor, improved toilet, improved water source, electricity in the house, radio, television, refrigerator, sofa, phone, and vehicle. The index was split into two wealth quintiles which were labeled as higher socio-economic status (higher SES and lower SES). These grouping are based on the SES of the participants in the sample, not the study population.

### Ethics

Ethics for this study was approved by the University of British Columbia (H17-02435), UCI (UCI-2022-39), and the Uganda National Council of Science and Technology (HS2420ES).

## RESULTS

**Table 1** provides the demographic information of the participants stratified by their socio-economic status. In all, 155 participants were classified in the higher SES category and 183 in lower SES category. Most notably, while only 17.4% (n=27) of the higher SES women reported being widowed, 37.7% (n=69) of the lower SES reported being widowed (p-value <0.001). Additionally, 74.9% (n=137) of the lower SES women had a primary school of less education compared to 46.5% (n=72) of the higher SES (p-value <0.001). In terms of stage, 15.8% (n=29) of the lower SES women were diagnosed with stage I compared to 6.5% (n=10) of the higher SES women (p-value <0.001).

**Table 1.**
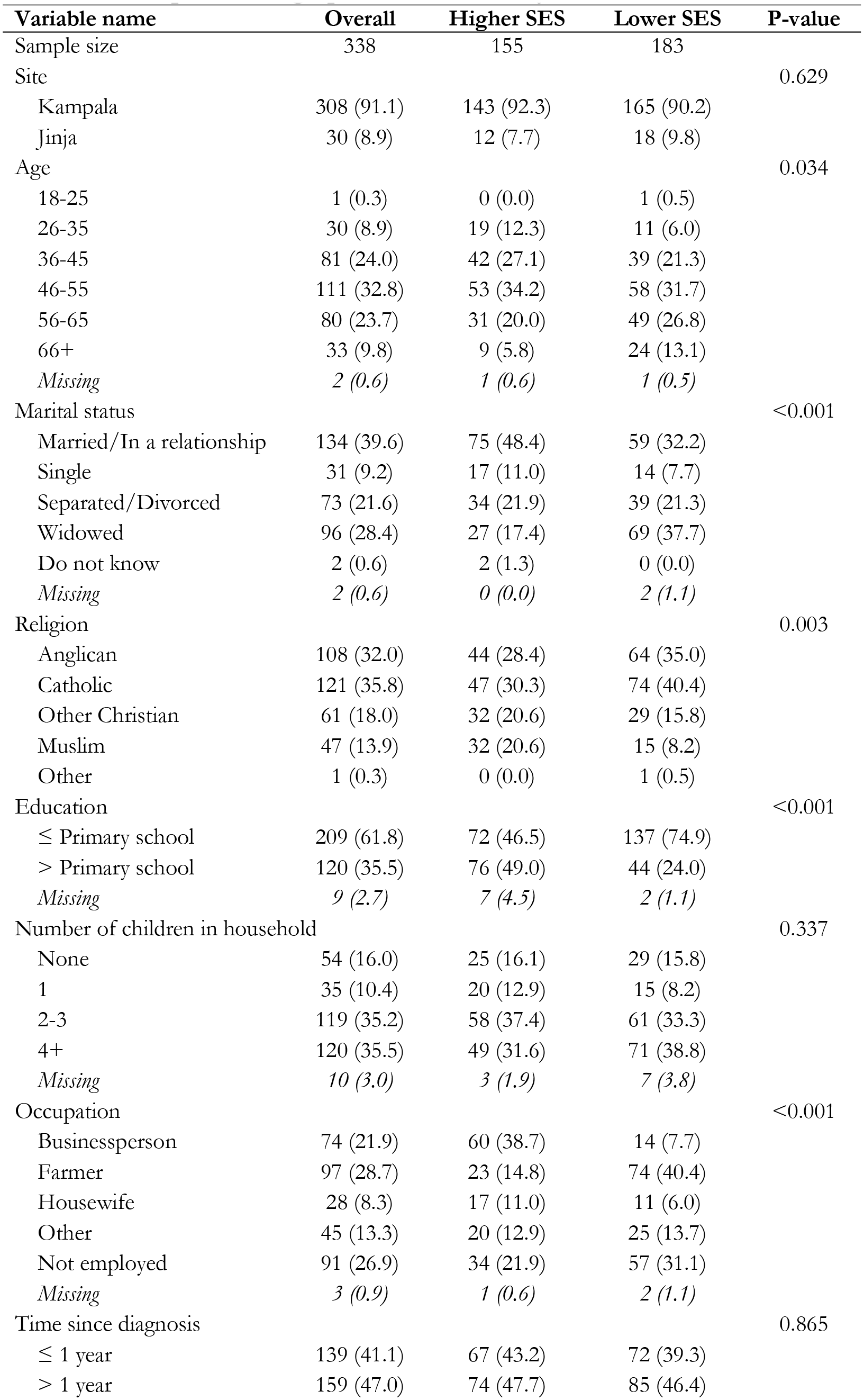
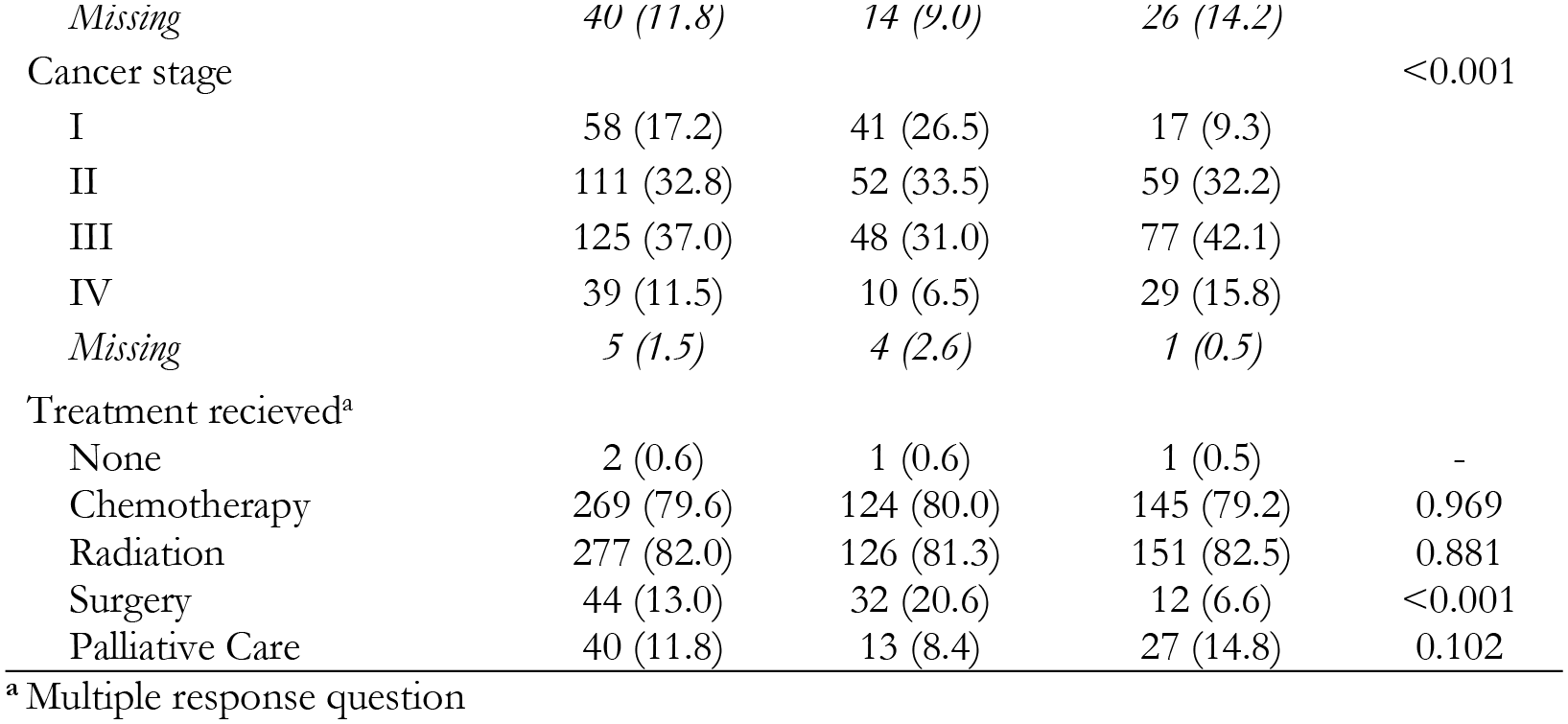
Participant demographic information by socio-economic status.

In all, 95.5% (n=148) of women from the higher SES and 92.3% (n= 169) of women from the lower SES reported that they paid out-of-pocket for at least one item related to their medical care (p=0. 3354). The most common items that women reported paying out-of-pocket for both SES groups were x-ray (higher SES n=129, 83.2%; lower SES n=157, 85.8%), scan (higher SES n=127, 81.9%; lower SES n=156, 85.2%), and radiation (higher SES n= 120, 77.4%; lower SES n=137, 74.9%). When asked how much they had spent, in total, on their cervical cancer treatment, 76 women of the higher SES and 93 women of the lower SES were able to provide an amount. The majority of the women in both groups reported between 500,001 to 2,000,000 UGX (between $130-530 CAD). **Table 2** reports on the indirect costs that women paid out-of-pocket for in the past month by SES. Over 90% of women from both SES groups reported paying for food and transportation costs related to their care.

**Table 2.**
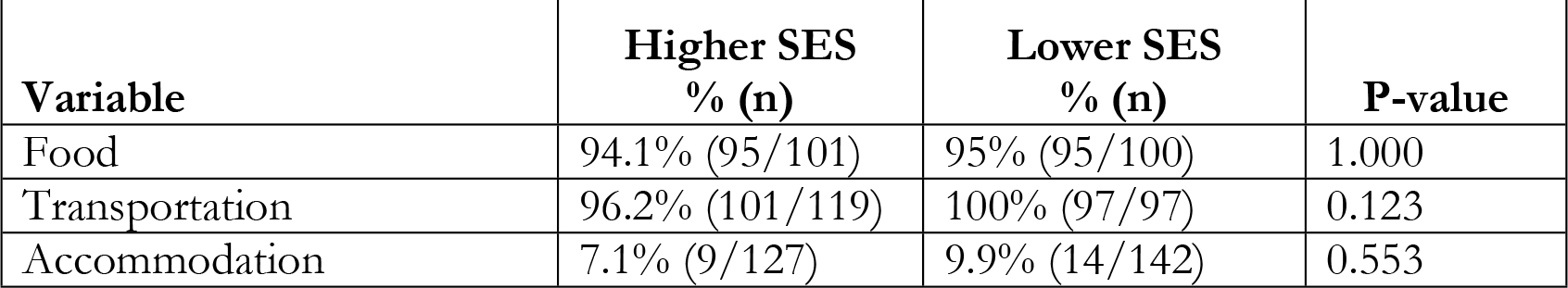
Indirect costs that women paid out-of-pocket for in the past month by SES.

Moreover, only nine (5.8%) women from the higher SES and six (3.3%) women from the lower SES reported that they stopped their cervical cancer treatment because they could not afford it. Among these 15 women, the most common treatments stopped due to cost were medications, biopsy, and radiation.

In all, only four (2.6%) women from the higher SES and six (3.3%) women from the lower SES reported that they received in-kind support. When asked about what kind of in-kind or financial support they needed while receiving treatment, 49 (31.6%) higher SES and 80 (43.7%) the lower SES said food, 50 (32.3%) higher SES and 73 (39.9%) lower SES said transportation, and 23 (14.8%) higher SES and 24 (13.1%) the lower SES said housing.

In all, 154 women from the higher SES and 181 from the lower SES reported on how they paid for their cancer care. Women of a lower SES were significantly more likely to report borrowing money (higher SES n=47, 30.5%; lower SES n=84, 46.4%; p-value =0.004) and selling possessions (higher SES n=47, 30.5%; lower SES n=90, 49.7%; p-value =0.006). On the other hand, women of a higher SES were statistically more likely to report not needing to seek out money to pay for their cervical cancer treatment (higher SES n=65, 42.2%; lower SES n=44, 24.3%; p-value =0.001).

In all, 44.3% of women in the higher SES (58/131) and 11.7% of women from the lower SES (14/120) indicated that they took time off work to attend the clinic today (p<0.001). There was no statistically significant difference between the two groups when asked if someone in their family either left their employment (higher SES n=66/140, 47.1%; lower SES n=62/147, 42.2%), reduced the number of hours worked (higher SES n=69/139, 49.6%; lower SES n= 71/147, 48.3%), or missed employment (higher SES n= 70/139, 50.4%; lower SES n= 74/153, 48.4%). However, women from the higher SES were more likely to report that someone in their household took time off of work to care for them (higher SES n=58/131, 44.3%; lower SES n=14/117, 14.0%; p-value < 0.001).

**Figure 1** highlights the change in time that women spent on unpaid labor since their cervical cancer diagnosis. Among the 109 higher SES women and 121 lower SES women who had a child under the age of 18, 31.2% of the higher SES group (n=34) and 29.8% (n=36) of the lower SES group reported that the amount of time they spent caring for their children decreased after their diagnosis. Furthermore, 31.5% of the higher SES group (n=34) and 26.4% (n=32) of the lower SES group reported that they decreased the amount of time that they helped their children with schoolwork.

**Figure 1.**
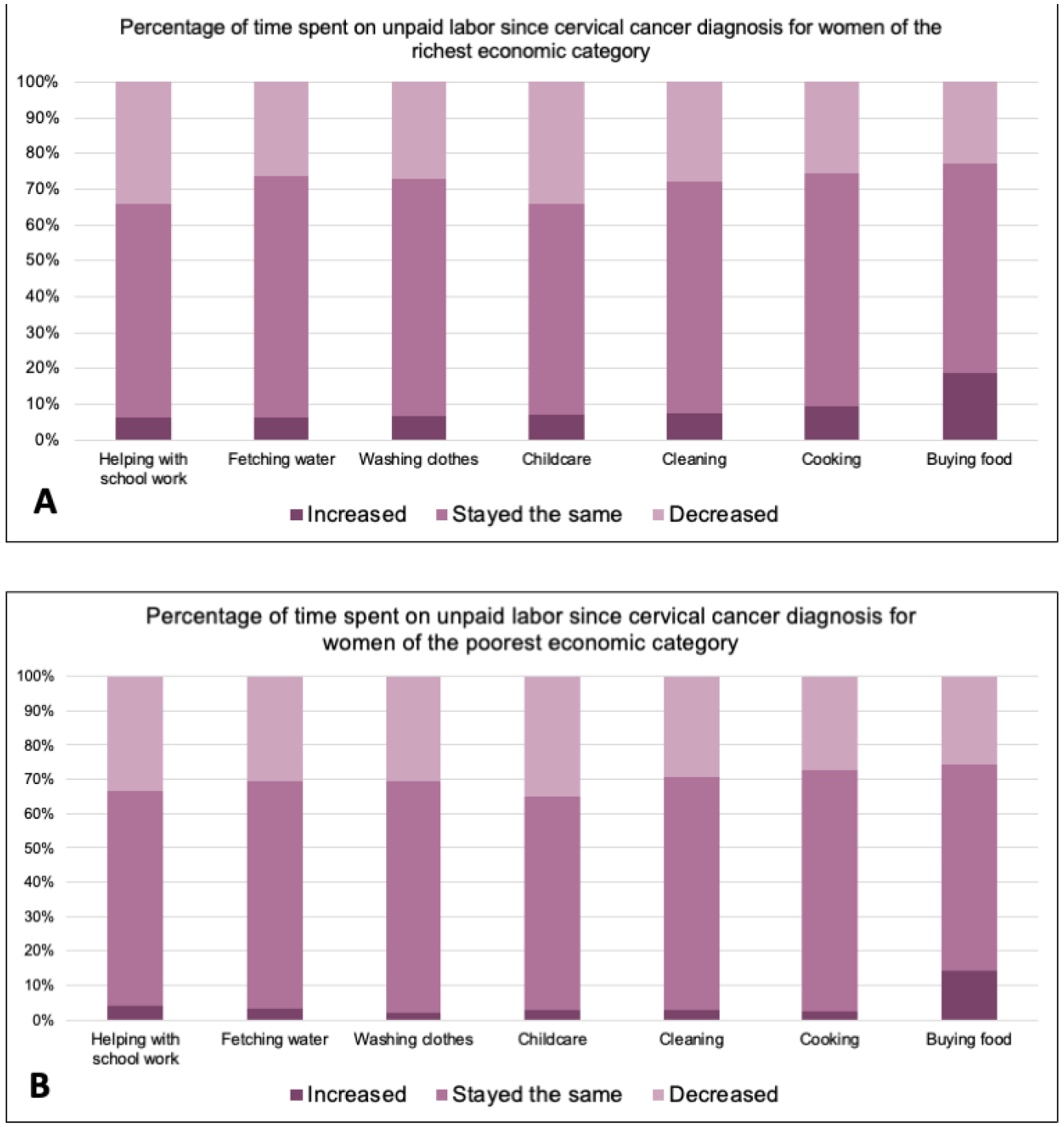
Changes in time spent on unpaid labor since the woman’s cervical cancer diagnosis stratified by higher SES (Figure A) and lower SES (Figure B).

## DISCUSSION

The results from the study show that women, regardless of their SES, incur out-of-pocket costs related to their cervical cancer treatment. The majority of women reported paying for common procedures such as a scan or radiation-costs which are not covered by the government. However, women in the lower SES group were more likely to report having to seek out funds to pay for treatment costs such as biopsy, chemotherapy, and laboratory tests. While these care costs are covered by the government, it is known that women are sometimes required to pay out of pocket for them. This means that while required costs are largely equal across socio-economic groups, there are incidences of inequity within the healthcare system. Moreover, we found that women from the higher SES group were more likely to have someone take time off of work to care for them. This can most likely be attributed to financial stability as families for a higher SES may have the economic support to allow for a member of their family to forgo wages to support them during their treatment.

An important finding from this analysis is that women from the lower SES group were more likely to be diagnosed with higher stage of cervical cancer. Most likely this is because they do not have regular, reliable access to screening. Common barriers to cervical cancer screening in Uganda include access to affordable, reliable transportation,[31-33] accessible, wide-spread, screening services and infrastructure,[33, 34] and available time to screen.[32] All of which can be exasperated by financial poverty. With that, it is not uncommon for women in Uganda to seek cervical cancer treatment not as a result of screening results, but because they are experiencing symptoms.[13] As such, these results highlight the urgent need to expand and improve the cervical cancer screening program in Uganda. One of the most demonstrated practical, efficient, and effective ways to achieve this is through a human papillomavirus (HPV)-based self-sampling program.[35] Although the HPV vaccine has a high uptake rate in Uganda (75% for the first dose)[36] and will reduce mortality in the long-term, there is still a need to screen immediately women for cervical cancer to prevent women from dying of this disease today. HPV-based self-sampling allows women to self-screen for cervical cancer on their own or under the care of a trained health worker.[37] Research in Uganda has shown that HPV-based self-sampling is an effective and acceptable way to screen women for cervical cancer.[38-40] Moreover, HPV-based self-sampling methods have shown to be the most cost-effective method, particularly when focused on increasing population coverage.[41, 42] Providing wide-spread, accessible HPV-based self-sampling would eliminate the economic burden of cervical cancer treatment on women and their families as it would prevent women from developing the disease.

We found in our analysis that although women are required to pay out-of-pocket for cervical cancer treatment, it does not lead them to stop care once they have started. In all, only 15 women from the study stopped a treatment because they could not afford it. Rather women cut costs or borrowed money to fund their cervical cancer care. However, we found, unsurprisingly, that women of the lower SES group were more likely to borrow money when compared to those of the higher SES group. This finding is aligned with a multitude of research in various healthcare fields on how individuals from the poorest economic status face the greatest financial strain when faced with healthcare costs.[43-46] This is important to highlight because it demonstrates that a cervical cancer diagnosis can have reverberating financial effects for those most vulnerable. These findings are further aligned with several other studies in Uganda which reported out-of-pocket healthcare expenditure at government funded hospitals.[5, 6, 47] For example, Anderson et al. reported in 2017 on surgical patients in southwestern Uganda and found that 12.5% to 33% of surgical patients were pushed into poverty from their medical costs.[6] With that, our findings demonstrate a clear need for additional economic support for financially vulnerable women in Uganda.

This analysis highlights that when women with children are diagnosed with cervical cancer and undergo treatment, they spend less time fulfilling childcare roles. In fact, we found that approximately one in three women from both SES groups with children under the age of 18 reported spending less time caring for their children and one in three from the higher SES group and one in four from the lower SES group spend less time helping their children with schoolwork since their diagnosis. This is important because globally, women are still expected to be the primary caregivers to children and, without them, children would be left without replacement support. While more research is needed, it cannot be assumed that this caregiving role is picked up by other family members such as husbands, mothers, or siblings. In fact, several research studies, including those in Ghana, Zambia, and Ethiopia, have reported in some incidences that male partners have abandoned their wives after their cervical cancer diagnosis.[48-50] With that, there needs to be a greater consideration as to how a woman’s cervical cancer diagnosis impacts the future generation as research has shown that children without mothers have poorer outcomes.[51-55]

A strength of this analysis is that it provides new and unique data on the individual financial burden of cervical cancer in Uganda. These novel findings can be used to make policy and care recommendations about cervical cancer in the country. Additionally, as we asked women questions pertaining to the wealth index and how much money they earned per month, this enabled us to validate the wealth index against the woman’s reported earnings. An important limitation of this study is that it only surveyed women who began treatment. This means that it excluded women who were diagnosed with cervical cancer but were unable to begin treatment due to financial barriers. This is important to highlight as it means these findings can only be taken in the context of those who were able to start treatment. Moreover, as this is a cross-sectional survey, we are unable to measure the direct change in a woman’s financial situation over time, which limits our understanding of the impact. These findings are additionally limited to individuals who attended government sponsored hospitals in Uganda. As such, these findings are limited in scope to patients who attend the UCI in Kampala or JRRH. With that, they cannot be applied to the entire population as they exclude individuals who attend private hospitals or seek care outside of the country.

This analysis highlights the economic burden that a cervical cancer diagnosis can have not only on women, but also their families. Although cancer care is largely subsidized in Uganda, findings from this study demonstrate that women often have to pay out-of-pocket for direct treatment costs. Moreover, many women, particularly those of lower SES backgrounds, must source funds to pay for their care. This puts a financial stress on women while they are undergoing treatment and also likely creates a prohibitive treatment barrier. With that, the Ugandan Ministry of Health should reflect on how healthcare costs can be financed more equitably. The Ministry of Health could consider universal health financing models such as a national health insurance program with tiered premium levels according to income. Revenues from a national health insurance program could help specifically fund the healthcare system, while ensuring that citizens continue to have access to care.

More funding for cancer treatment programs through revenue such as a health insurance program would help offset the necessary costs for individuals undergoing treatment. In addition to this, our study supports the expansion of satellite clinics into more communities to help alleviate transportation, housing, and food costs for women travelling to the clinic, as this was a major source of out-of-pocket costs. This in turn would help prevent families from experiencing financial catastrophe and reduce the stress of seeking care. Additional education to destigmatize cervical cancer and traditional gender roles would help support mother’s undergoing treatment. Most importantly though, the Ugandan Ministry of Health should consider expanding the screening and vaccination program for cervical cancer in order to prevent women from being diagnosed and dying of this preventable disease.

## Data Availability

Data cannot be shared publicly due to ethical restrictions, as participants did not provide consent at the time of data collection for data to be shared publicly. Anonymized minimal data set will be made available by request for researchers who meet the criteria for access to confidential data and approval by the UBC Research Ethics Board (contact via)

## Acknowledgements

We would like to thank the participants for sharing their time. We would also like to acknowledge Shamim Nakyazze for her assistance with study coordination.

